# Fingerstick blood assay maps real-world NAD+ disparity across gender and age

**DOI:** 10.1101/2023.02.07.23285529

**Authors:** Pei Wang, Meiting Chen, Yaying Hou, Jun Luan, Ruili Liu, Liuqing Chen, Min Hu, Qiuliyang Yu

## Abstract

NAD^+^ level has been associated with various age-related diseases and its pharmacological modulation emerges as a potential approach for aging intervention. But human NAD^+^ landscape exhibits large heterogeneity, and the lack of rapid, low-cost assays limits the establishment of NAD^+^ baseline and the development of personalized therapies, especially for those with poor responses towards conventional NAD^+^ supplementation. Here, we developed an automated NAD^+^ analyzer for the rapid measurement of NAD^+^ with 5 μL of capillary blood using a recombinant bioluminescent sensor protein and an automated optical reader. The minimal invasiveness of the assay allowed a frequent and decentralized mapping of real-world NAD^+^ dynamics. We showed that sports and NMN supplementation can increase blood NAD^+^ levels and that male on average has higher NAD^+^ than female before the age of 50. We further revealed the long-term stability of human NAD^+^ baseline over 100 days and identified the major real-world NAD^+^-modulating behaviors.

## Introduction

NAD^+^ is an essential substrate for numerous redox reactions and regulatory proteins including poly(ADP-ribose) polymerase (PARPs), Sirtuins, CD38/157 and SARMs, which play important roles in DNA repair, protein deacetylation, immune response, and other fundamental cellular processes^1–5^. The decline of NAD^+^ content in blood and saliva has been associated with aging and age-related symptoms such as frailty^4,6^, rheumatoid arthritis^7,8^, and heart failure^9,10^ at old age. In addition, oral administration of NAD^+^ precursors as a potential intervention strategy of age-related symptoms has been demonstrated in clinical trials to improve body NAD^+^ levels^5,11–15^, muscle functions and skeletal muscle insulin sensitivity and signaling^16,17^, establishing NAD^+^ level as an emerging indicator of healthy aging.

However, as considerable disparity exists in both the geno- and phenotype of human aging, the blood NAD^+^ landscapes of different people and aging processes are yet to be fully characterized^18,19^. One major limitation for mapping the NAD^+^ landscape at the point of care is its quantification method. Conventional NAD^+^ measurement relies on High-Performance Liquid Chromatography (HPLC)^20,21^, Liquid Chromatography-Mass Spectrometry (LC-MS)^22–24^, enzyme cycling assays^25^ or Nuclear Magnetic Resonance (NMR)^26–28^, which requires centralized laboratories and multi-step sample preparations. Recently developed semisynthetic bioluminescent sensors offered a solution for measuring NAD^+^ at point-of-care^29^. But this sensor relies on a synthetic chemical tether that requires multi-step synthesis with relatively low yield, limiting its mass production. Hence, more cost-effective and easy-to-produce protein sensors should make the assay much more available and affordable for the large-scale, real-world mapping of NAD^+^ dynamics.

Here we developed an NAD^+^ assay using an *Escherichia coli* (*E*.*coli*)-produced^30^, fully genetically encoded NAD^+^ sensor and a simple, automated desk-top reader for measuring NAD^+^ levels in venous and capillary blood, as well as in saliva with 5 μL of the sample. We used this assay in a placebo-controlled, double-blind clinical trial to assess the effect of NMN and sports. We showed that oral NMN supplementation and sport can significantly increase blood NAD^+^ levels. In addition, the high-level agreement between the sensor and HPLC-MS established the sensor as a viable NAD^+^ quantification method. The assay further mapped the NAD^+^ levels in capillary blood and revealed a marked NAD^+^ disparity across gender and age. Moreover, the sensor made long-term and frequent NAD^+^ monitoring easily available due to the minimal invasiveness of the capillary sampling, with which we mapped the speculated NAD^+^ Circadian rhythm, short-term pharmacodynamics of oral NMN administration, and the long-term real-world NAD^+^ baseline in human.

## Results

### Measuring blood NAD^+^ using recombinant bioluminescent sensor protein

For achieving low-cost and rapid NAD^+^ measurement in clinical samples, we developed an NAD^+^ assay using a simple, automated reader and a fully genetically encoded bioluminescent NAD^+^ sensor protein named NS-Goji 1.3. The recombinantly expressed sensor contains an engineered NAD^+^-binding domain, a circularly permuted luciferase cpNLuc and a red fluorescent protein (RFP) mScarlet-I (Fig. 1A). NS-Goji 1.3 undergoes considerable conformational changes upon NAD^+^ binding and shortens the distance between cpNLuc and mScarlet-I, thus affecting the BRET efficiency between the pair (Fig. 1B). The highly specific NAD^+^-dependent shift of the sensor’s emission intensity at 460 nm and 580 nm is used for quantifying the NAD^+^ levels in the samples (Fig. 1C). To automatize the assay, we developed a reader that integrates a single-channel pipette robot, a photon-detector, and a reagent-containing test strip. As the *E. Coli* expression system makes the sensor production cost-effective and scalable, the NAD^+^ sensor is easily available for large-scale sample analysis. In parallel, an HPLC-MS quantification procedure for NAD^+^ has been developed as the reference method in which NAD^+^ was identified by ion pair of 662.0/539.7 and 80.0 m/z and quantified by the intensity of ion 662.0 m/z (Fig. 1D, 1E, and 1F).

**Figure 1.**
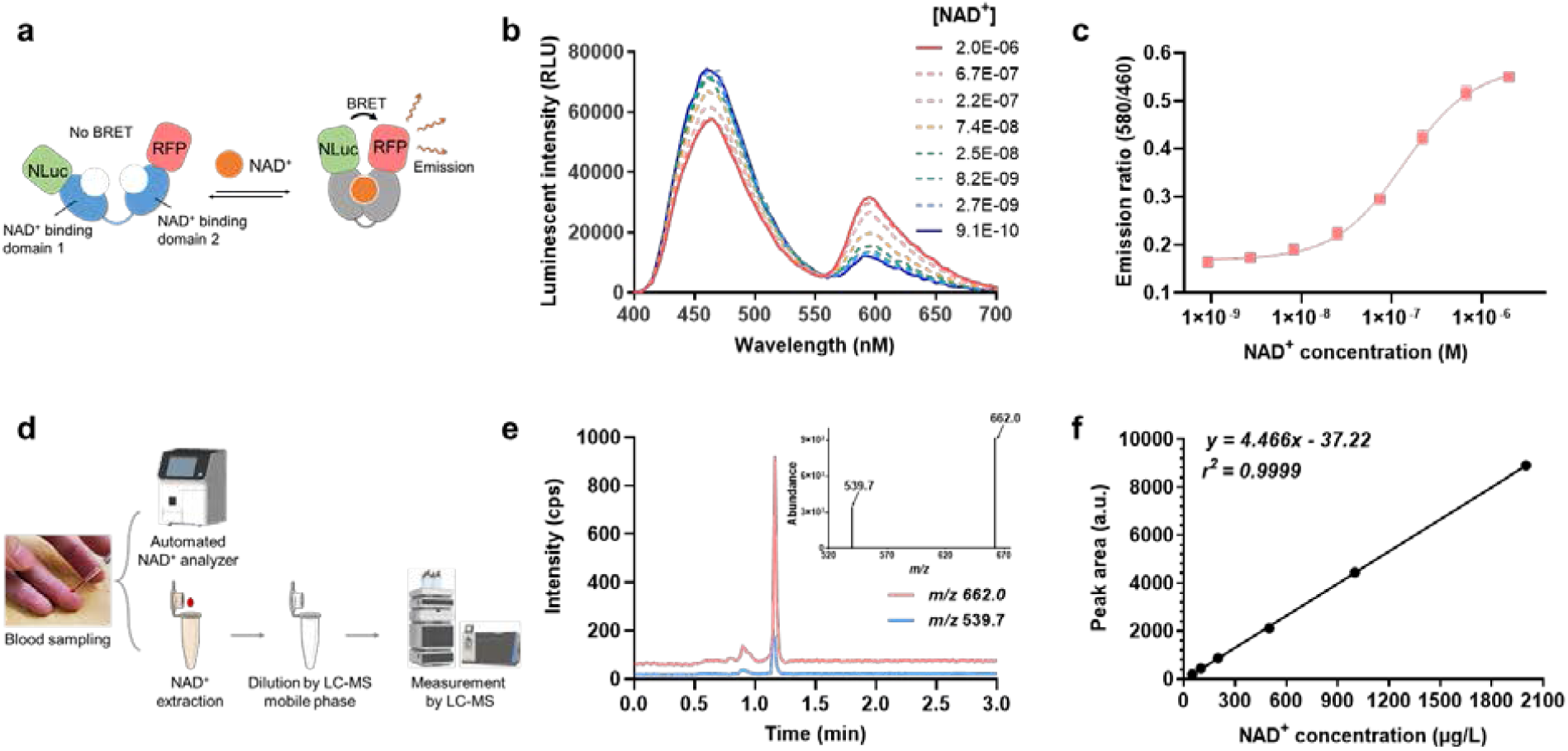
Quantification of whole blood NAD^+^ by sensor and LC-MS. **(a)** Sensing mechanism of the recombinant bioluminescent NAD^+^ sensor protein NS-Goji 1.3: The sensor changes conformation as a function of NAD^+^ concentration, leading to changes in the BRET efficiency between luciferase NLuc and RFP m-Scarlet I. **(b)** Sensor’s emission spectra at various NAD^+^ concentrations. **(c)** Ratios between sensor’s emission intensities at 580 nm and 460 nm plotted against various NAD^+^ concentrations. Values are given as mean ± SD of three independent measurements. **(d)** Sample preparation procedure for NAD^+^ analyzer and HPLC-MS quantification. **(e)** HPLC-MS retention time and ionization intensity of ions pairs for NAD^+^ identification and quantification. **(f)** Calibration curve for NAD^+^ measurement by HPLC-MS.

### Sports and oral NMN significantly increase venous NAD^+^

Using the analyzer, we quantified the NAD^+^ levels in venous blood obtained from clinical studies to evaluate the effect of NMN supplementation on subjects between 55-70 years of age (Table S1). The double-blinded clinical test compared one placebo group and two treatment groups using different dosages of NMN (500 mg/day and 1000 mg/day) (Fig. 2A). After 30 days of daily oral administration, venous blood was collected for assessing whole-blood NAD^+^ content using both the sensor and HPLC-MS.

**Figure 2.**
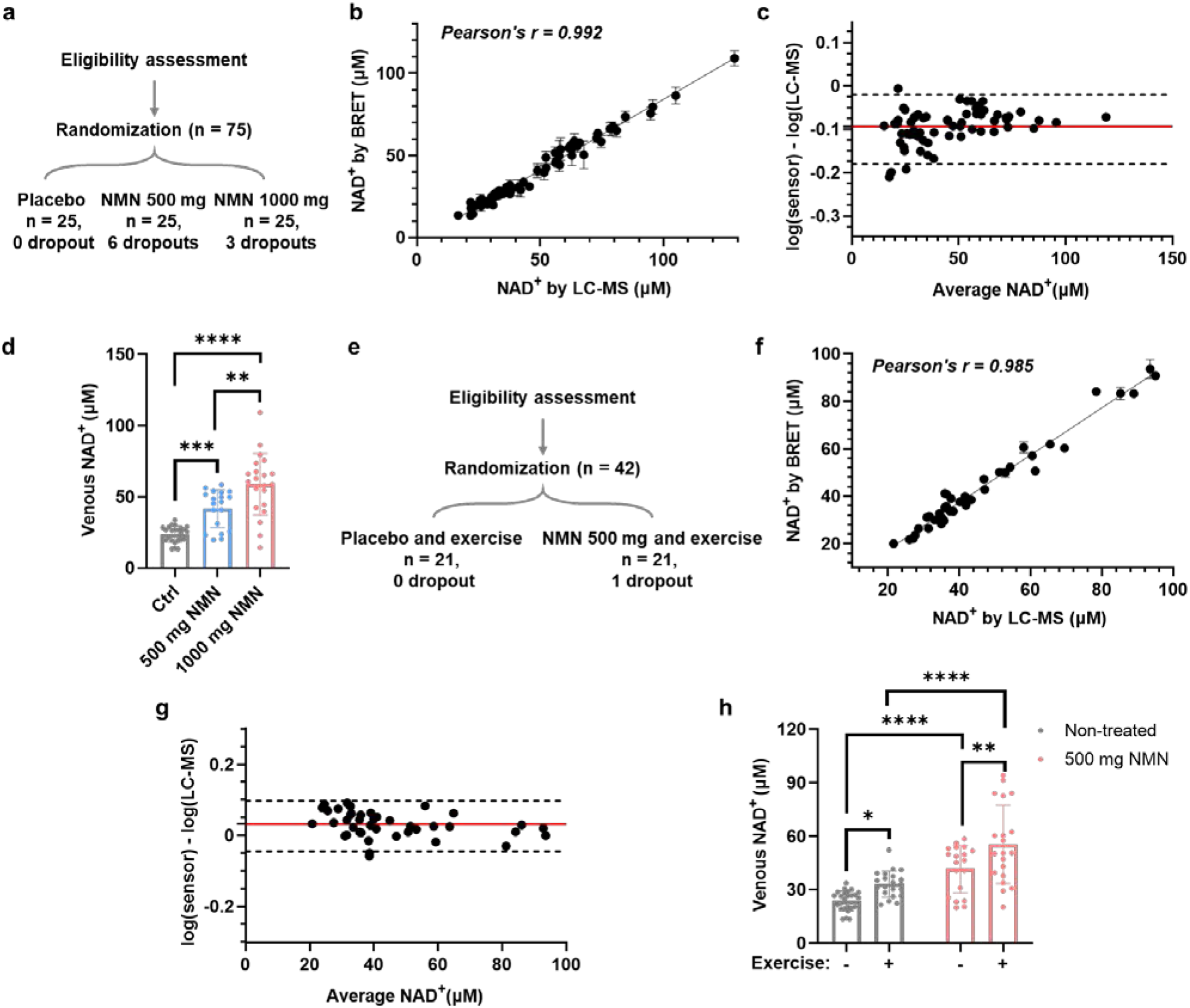
Measurement of venous NAD^+^ levels in clinical studies of NMN supplementation and sport. **(a)** Study design for evaluating effects of oral NMN supplementation. **(b)** Comparison of NAD^+^ levels measured by sensor protein and HPLC-MS. **(c)** Bland-Altman analysis for venous NAD^+^ measured by sensor and HPLC-MS. **(d)** Effect of oral NMN supplementation on venous NAD^+^ level. Daily supplementation of 500 mg (n = 19) and 1000 mg NMN (n = 22) for 1 month significantly increased the venous NAD^+^ concentration compared to the placebo group (n = 25). **(e)** Study design for evaluating effects of oral NMN supplementation in combination with moderate level of sport. **(f)** Comparison of NAD^+^ levels measured by sensor protein and HPLC-MS. **(g)** Bland-Altman analysis for venous NAD^+^ measured by sensor and HPLC-MS. **(h)** Effect of sport with oral NMN supplementation (n = 20) and sport alone (n = 21) on venous NAD^+^ level. Sports significantly increased venous NAD^+^ levels in both placebo and NMN-treated groups. In (b) and (f), values are given as mean ± SD of three independent measurements. In (d) and (h), error bars represent SD of the respective group. Significance was determined using one-way ANOVA analysis for (d) and two-way ANOVA analysis for (h), * *p* < 0.05, ** *p* < 0.01, *** *p* < 0.001, **** *p* < 0.0001.

The measurement revealed that both dosages of NMN significantly increased the venous NAD^+^ in a dose-dependent manner. The recorded average NAD^+^ level was 23.8 ± 5.5 μM in the placebo group, 41.7 ± 13.0 μM in the 500 mg/day group and 58.8 ± 21.1 μM in the 1000 mg/day group. Notably, the variation of NAD^+^ level in the placebo group is smaller compared to that of the two treatment groups. The group treated with 1000 mg/day oral NMN featured a wide distribution of the NAD^+^ levels (SD = 21.1), indicating the possible non-responses towards the NMN administration. In addition, the comparison between NS-Goji 1.3 and HPLC-MS demonstrated a good agreement between the two methods with Pearson’s r = 0.992 (Fig. 2B, and 2C). In the subsequent clinical study, we evaluated the effect of sports on the venous NAD^+^ levels of the placebo- and NMN-treated (500 mg/day) subjects for 30 days (Fig. 2E, Table S2). NS-Goji 1.3 achieved a similar level of quantification compared to HPLC-MS for measuring venous NAD^+^ with Pearson’s r = 0.985 (Fig. 2F, and 2G). The blood NAD^+^ test showed that the moderate-intensity physical exercise significantly increased the NAD^+^ levels of both the placebo- and NMN-treated subjects compared to the control, indicating frequent exercise as a potential strategy to maintain NAD^+^ levels and to enhance the response towards NAD^+^ supplementation (Fig. 2H).

In addition to measuring NAD^+^ in blood, we explored the possibility of NAD^+^ measurement in saliva. Recent work for identifying salivary aging biomarkers revealed that salivary NAD^+^ decreases with age^31^. We hence tested if the bioluminescent sensor could quantify NAD^+^ in salivary samples. The analysis performed in parallel with HPLC-MS demonstrated that the bioluminescent sensor is capable of testing salivary NAD^+^ with Pearson’s r = 0.999 (Fig. S1). Overall, we showed that NS-Goji 1.3 sensor and the automated analyzer are viable tools for scalable and low-cost measurements of NAD^+^. The clinical study further demonstrated that regular sports enhanced the NAD^+^ in both placebo- and NMN-treated subjects.

### Fingerstick blood NAD^+^ survey reveals gender-associated differences in NAD^+^ levels

For conventional NAD^+^ quantifications, venipuncture limits the frequency of NAD^+^ sampling due to invasiveness. By contrast, collecting fingerstick capillary blood by disposable lancet (Fig. 3A) would largely facilitate the dissemination of NAD^+^ measurement, which is now made possible by the bioluminescent sensor. To evaluate if fingertip blood is a representative sample source for quantifying NAD^+^, we compared the venous and fingerstick capillary blood obtained in parallel from 13 volunteers within 30 min. The NAD^+^ measured from the two sample types showed a good correlation (Pearson’s r = 0.987) and are within ± 15% error (Fig. 3B and Fig. S2). In this test, NS-Goji 1.3 measurement typically requires 5 μL of fingertip blood, while by contrast, a single venipuncture collection requires several milliliters of sample.

**Figure 3.**
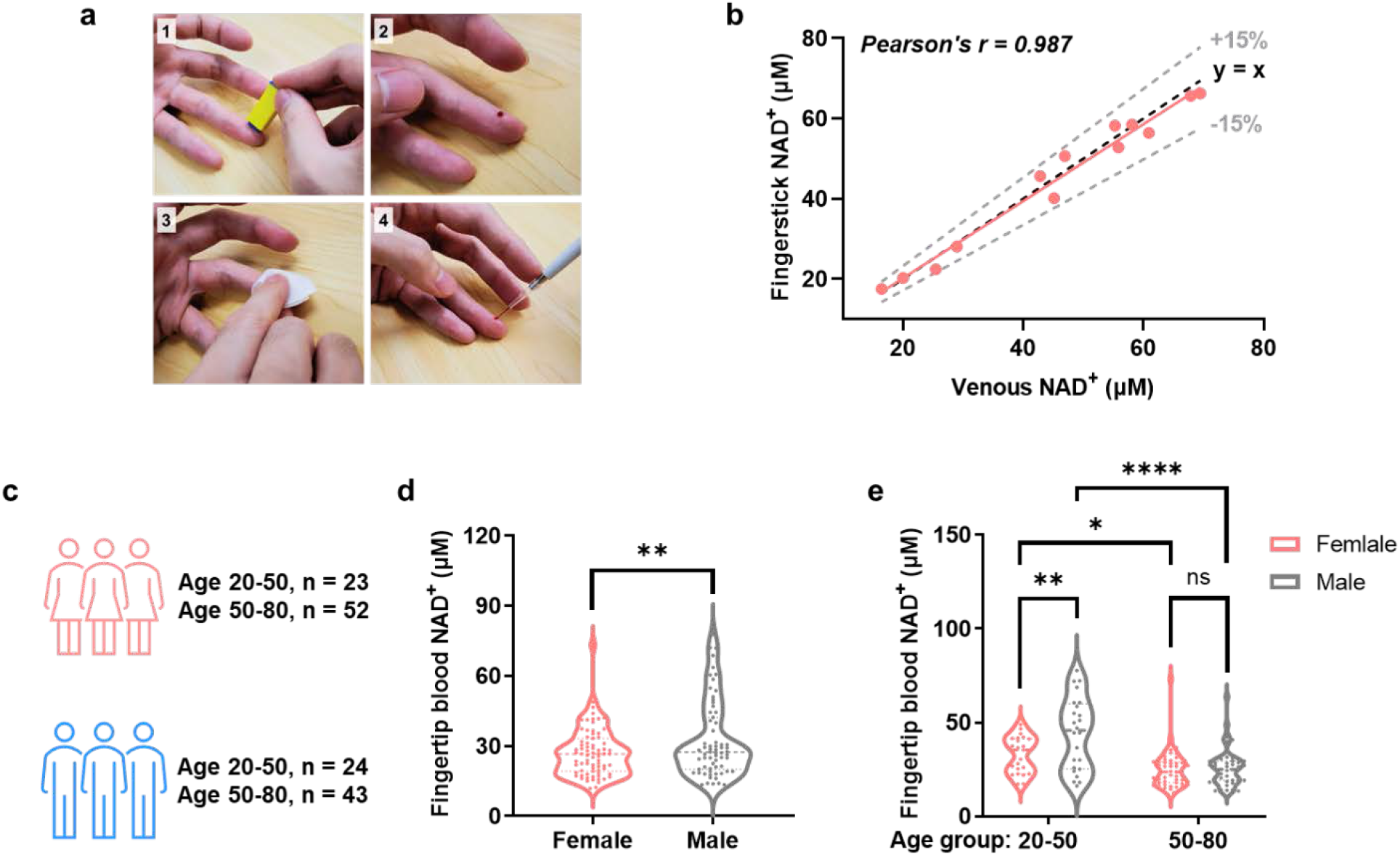
Fingertip blood NAD^+^ measurement for mapping NAD^+^ disparity. **(a)** Procedures of taking fingerstick capillary blood for NAD^+^ measurement. **(b)** Comparison between NAD^+^ levels measured from fingertip and venous blood. The Black dashed line indicates y = x, and the gray dashed line indicates ± 15% deviation from y = x. **(c)** Study design for mapping fingertip blood NAD^+^ levels for healthy volunteers with different age and gender. **(d)** Female participants (n = 75) showed a significantly lower NAD^+^ level than males (n = 67) in this study. **(e)** 50-85 age group has significantly lower NAD^+^ levels than 18-50 age group for both male and female. The male has higher NAD^+^ level than female for 18-50 age group but not for the 50-85 age group. Significance was determined using t-test for (d) and two-way ANOVA analysis for (e), * *p* < 0.05, ** *p* < 0.01, **** *p* < 0.0001.

Using this fingerstick blood NAD^+^ assay, we further mapped the NAD^+^ levels of NAD^+^ supplement-naïve people across different age groups (Table S3). The cross-gender comparison indicated that females have on average a lower NAD^+^ level (27.2 μM) compared to males (32.5 μM, Fig. 3C). The survey further revealed that participants between 18-50 years of age have significantly higher NAD^+^ content than those aged between 50-85 (Fig. 3D) for both genders. Notably, the age-related NAD^+^ decrease is more pronounced for males with the average NAD^+^ level decreased from 44.2 μM to 25.9 μM (P < 0.0001). While for females, the NAD^+^ content is decreased by a lesser extent from 32.7 μM to 24.8 μM with P < 0.05. The gender-related NAD^+^ disparity is more pronounced for participants younger than 50 years old (Fig. 3D). But no significant gender-related differences were observed for participants aged over 50 (Fig. 3D). Interestingly, a few participants have particularly high NAD^+^ levels for the respective age group, indicating the existence of outliers whose NAD^+^ metabolism is markedly different. Further studies to identify the genetic and/or acquired factors for this high NAD^+^ level should provide more insight for developing new NAD^+^ modulating strategies.

### Measuring real-world NAD^+^ dynamics using minimal capillary blood samples

NAD^+^ metabolism plays a central role in Circadian rhythm. However, the hypothetical human NAD^+^ differences between day and night have not been recorded due to the difficulties of collecting venous blood after midnight. As fingertip blood assay is easily applicable at the bedside, we collected and measured the capillary blood NAD^+^ at 4 am and 10 am from n = 7 subjects (Fig. 4A). The measurement showed that the capillary NAD^+^ levels did not change significantly between 4 am at 10 am (P = 0.0735, Fig. 4B). To evaluate the potential fluctuation in NAD^+^ during the day, we further accessed the capillary NAD^+^ levels after oral administration of 300 mg of NMN during the day starting from 10 am. The NAD^+^ level significantly increased 60 min after the NMN administration and returned quickly to the basal level at 120 min (Fig. 4C). Apart from the short NMN-induced perturbation, no significant changes were observed during the day, demonstrating the fast metabolism of NMN and the strong homeostasis of the NAD^+^ level. To further map the NAD^+^ fluctuations in the real world over long terms, we measured the NAD^+^ levels of n = 6 subjects twice a week over 100 days and recorded major changes in dietary, sportive, and sleeping habits (Fig. 4D). The result showed that NAD^+^ levels were mostly stable during the 100-day monitoring (Fig. 4E&F), except for one participant who spontaneously started NMN administration which induced a notable increase in the capillary NAD^+^ level (from 32.9 to 47.3 with p < 0.0001). One subject exercised regularly (11 out of 25 days on which NAD^+^ was measured), and the recorded NAD^+^ levels on days with sport were significantly higher than days without. However, the sport-induced NAD^+^ increase was not observed for the subject with a low sport frequency (3 out of 24 days of NAD^+^ measurement). Other recorded events such as sleep deprivation, menstruation, oral administration of phosphatidylcholine (PPC) and silibinin did not significantly affect the capillary NAD^+^ level (Fig. 4G).

**Figure 4.**
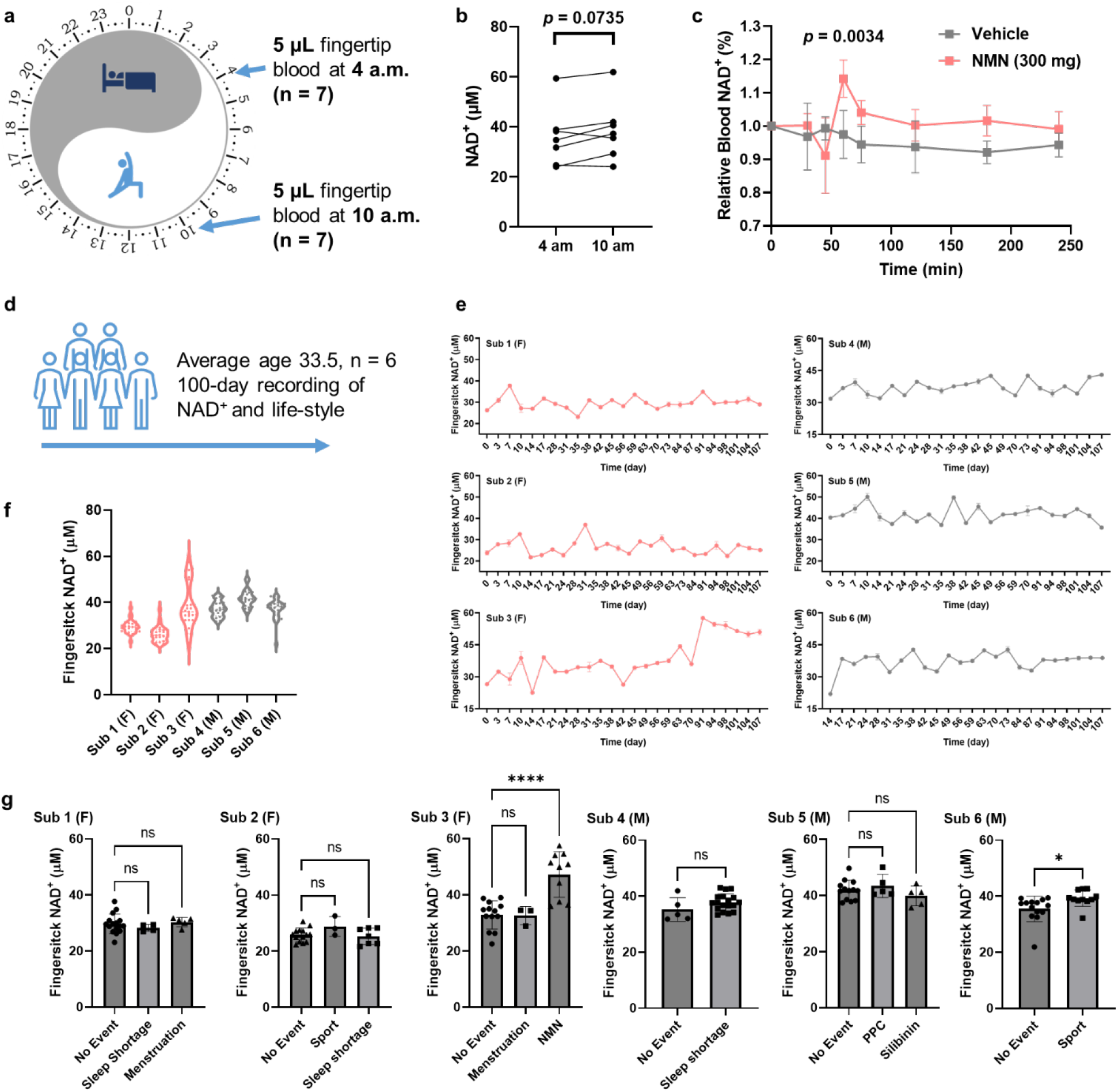
Monitoring real-world NAD^+^ dynamics using capillary blood. **(a)** Study design for comparing capillary NAD^+^ levels at 4 and 10 a.m. **(b)** No significant difference in NAD^+^ level was observed for capillary blood samples obtained at 4 and 10 a.m. (n = 7, *p* = 0.0735), though the average NAD^+^ level increased from 36.00 μM to 38.69 μM. **(c)** Oral administration of 300 mg NMN induced short-term capillary NAD^+^ spike with significant increases recorded at 60 min (n = 5, *p* = 0.0034). **(d)** Scheme for the long-term monitoring of NAD^+^ levels. Events including sport, sleep shortage, medication, menstruation, and dietary supplementation were recorded through questionnaire. **(e)** NAD^+^ dynamics recorded for n = 6 participants over three months. F and M indicates female and male subjects respectively. **(f)** Distribution of fingerstick NAD^+^ levels during the long-term monitoring. **(g)** Effects of events on NAD^+^ levels for each subject. NMN and sport significantly increased NAD^+^ levels for sub 3 and 6 respectively. Only days with single recorded event were considered, while days with more than one recorded event were excluded from the analysis. Significance was determined using t-test for (c) and sub 4 & 6 in (g). Two-way ANOVA analysis was used for significance determination for sub 1, 2, 3 and 5 in (g), * *p* < 0.05, **** *p* < 0.0001.

Taking together, the capillary blood NAD^+^ measurement revealed significant NAD^+^ disparity across gender and age. The sensor also facilitated the real-world observation of NAD^+^ modulation by dietary supplements and sports. We further demonstrated that individual NAD^+^ levels are relatively stable over a long period of time and continuous oral NMN administration and frequent sport can boost NAD^+^ levels. This easy-to-implement NAD^+^ test for blood and saliva provided a useful tool for monitoring the effect of various NAD^+^ modulating strategies at the point-of-care.

## Discussion

Blood NAD^+^ level is becoming an important physiological parameter for evaluating NAD^+^ boosting strategies. Several previously reported clinical studies of NAD^+^ supplementation established that blood NAD^+^ content decreases with the progression of age-related diseases such as mitochondrial myopathy and rheumatoid arthritis^7,32^. In addition, previous studies showed that oral supplementation of NA^12^, NR^33–35^ or NMN^17,36^ can increase the blood NAD^+^ level and lead to beneficial outcomes. However, the conventional quantification based on HPLC-MS or enzymatic cycling assays are time- and labor-intensive, hence limiting the up-scale and generalization of the NAD^+^ blood test. Here, we demonstrate that the low-cost and recombinantly expressed bioluminescent NAD^+^ sensor protein enables easy-to-implement assays for analyzing microliters of clinical samples with a good coherence compared to HPLC-MS.

The two doses (500 mg and 1000 mg per day) of oral NMN supplementation evaluated in this work can induce a significant increase in blood NAD^+^ level. However, considerable variation exists for the group supplemented with 1000 mg NMN per day, indicating the heterogeneity of NMN responses among the subjects. Recent work on the NAD^+^ flux indicated an age-related increase of CD38 and the resulting NAD^+^ consumption, contributing to the NAD^+^ decline during aging^37,38^. The existence of NMN non-responders may be due to the hyper-activated NAD^+^ consumption pathways such as CD38 which degrades NMN and NAD^+^ as substrates. More detailed studies would be necessary to better characterize these subgroups and to enhance the response rate of NAD^+^ supplementation. To achieve better supplementation outcomes for the non-responders, NAD^+^ precursors and modulators of its consumption pathways may be used in combination, and the convenient NAD^+^ blood monitoring can help to evaluate the effect of the combo therapies. Using the NAD^+^ sensor, we also demonstrated that regular sports could increase the NAD^+^ basal level and enhance the NMN response among the participants, providing precursor-independent alternatives to NAD^+^ modulation.

As considerable disparity exists during human aging, mapping and characterizing various subgroups of the aging phenotype is key for achieving personalized aging intervention. However, collecting large numbers of clinical samples has been challenging for mapping the NAD^+^ metabolism among different populations. Here, the bioluminescent sensor enabled an easy-to-implement assay for quantifying NAD^+^ using fingertip blood at a much-reduced cost compared to HPLC-MS. The NAD^+^ survey revealed a gender-related NAD^+^ disparity which is especially pronounced before the age of 50. The survey also identified NAD^+^ supplement-naive individuals with particularly high NAD^+^ levels compared to their peers. As this assay is minimally invasive and cost-effective, future surveys at larger scales covering different geographic and ethnic groups may identify more naturally occurring cases with high NAD^+^ levels. Further studies to elucidate the underlying mechanisms should provide new exploitable strategies for NAD^+^ modulation and aging intervention.

Furthermore, we established that human NAD^+^ is generally stable during the day and night, as well as over long periods of time. However, NMN administration can induce notable spikes in NAD^+^ and may shift the original NAD^+^ homeostasis. The elevated NAD^+^ level can be maintained by the NMN administration, but a safe and effective NAD^+^ window induced by such intervention is yet to be determined. In addition, we observed that regular sports with moderate intensity readily increases the NAD^+^ levels though by a lesser extent than the NMN administration. Despite that no short-term adverse events were observed in neither our study nor other reported clinical studies of NMN administration, establishing therapeutic windows for NAD^+^ supplementation should be important for the long-term management of aging-related symptoms.

Several limitations should be considered in our study. First, we recruited 76 females and 67 males for assessing the age-related NAD^+^ disparity. Even though the sample size is sufficient for evaluating the age-related decline of NAD^+^, having more participants would be beneficial for detecting the potential NAD^+^ differences between more refined age groups. Second, our longitudinal analysis showed that sports and NMN increases NAD^+^ for the tested individuals, but more data points and larger sample sizes will help to identify other NAD^+^-modulating behaviors and establish new sportive or dietary regimes to enhance NAD^+^ during physiological aging. The analysis excluded the data points with more than one event recorded on the same day and did not analyze the combined effects of multiple events on the NAD^+^ level due to the limited data set. Combining NAD^+^ levels with more comprehensive and continuous data obtained by health-monitoring wearables such as smartwatches will provide deeper insights into how different daily behaviors and habits can influence NAD^+^ metabolism and provide new approaches for its active modulation. Third, the NAD^+^ survey indicated the existence of individuals with notably high NAD^+^ levels. However, we could not yet characterize the genetic or metabolic features of the individuals within the scope of this study. But the cost-effective NAD^+^ survey will enable the mapping of NAD^+^ at larger scales and lead to future investigations into the underlying mechanisms.

Overall, we demonstrated a low-cost and easy-to-use assay using recombinant sensor protein and an automated optical reader for measuring fingertip blood NAD^+^ and revealed how parameters such as NAD^+^ supplementation, sports, gender, age, Circadian rhythm etc., may contribute to the NAD^+^ disparity amongst people. The widely observed differences in human aging underline the necessity of developing easily available, minimally invasive, and low-cost sensing tools for measuring aging biomarkers to identify meaningful subgroups of the aging phenotype and to empower personalized aging interventions through the quantitative monitoring of NAD^+^-modulating regimes.

## Methods

### Clinical subjects and samples

Study protocol for evaluating the effect of oral NMN supplementation and sports on human subjects have been reviewed and approved by the ethics committee of Guangzhou Sport University (ethics approval number: 2020 LCLL-003) in accordance with the Declaration of Helsinki. First, the study evaluates how different doses of NMN affect the blood NAD^+^ level. Then, the study compares the effect of sports with and without NMN supplementation on the blood NAD^+^. The study recruited participants between 55-70 years of age. The exclusion criteria are 1) having hepatic and/or renal dysfunction, infectious diseases, cardiac dysfunction, and/or spinal dysfunction, 2) having chest pain or shortness of breath during sport, 3) having more than 3% changes of body weight within 2 months before the study, 4) having previously taken nicotinic acid, nicotinamide mononucleotide, nicotinamide, or other vitamins B3-related dietary supplements, 5) taking coffee or caffeine-containing drinks, 6) during pregnancy or lactation, 7) being mentally unfit to participate in the study. For the sport intervention, subjects participated in centralized aerobic sport for 40-60 min, three times per week. For the first two weeks, the sport intensity was maintained between 40% to 59% of the heart rate reserve (HRR), then sport intensity was increased to 60% to 85% of the HRR. In addition, subjects participated in resistance training 3 times per week. For the dietary supplementation, subjects were grouped and tested in a double-blinded, placebo-controlled manner with three groups taking the placebo, 500 mg/day NMN, and 1000 mg/day NMN.

The study protocol for evaluating fingertip blood NAD^+^ levels of human subjects have been reviewed and approved by the Institutional Review Board of Shenzhen Institute of Advanced Technology, Chinese Academy of Sciences (ethics approval number: SIAT-IRB-210915-H0575) in accordance with the Declaration of Helsinki. Fingerstick sampling was performed to collect 5 μL of capillary blood using disposable fingerstick lancets (STERiLANCE Medical Inc.) for NAD^+^ measurement. The blood sampling time, administration of medication and/or dietary supplements, physical exercise, sleep quality and menstruation have been recorded for data analysis with written consent of participants. The exclusion criteria are 1) being younger than 18 years of age, and 2) having hemophobia or other symptoms hindering the fingerstick blood sampling.

### Sensor preparation

The gene of the sensor NS-Goji 1.3 was constructed in a pET15b (+) vector for recombinant expression in *E. coli* (DE3). The *E. coli* (DE3) was cultured 1 L LB medium containing 50 μg/mL ampicillin at 37 °C with a shaking rate of 220 rpm to reach an optical density of 0.6-0.8 at 600 nm. Then 0.5 mM isopropyl β-D-thiogalactopyranoside (IPTG) was added into the medium to induce sensor expression at 16 °C for 16 hours. The *E. coli* (DE3) was collected by centrifugation at 8000 rpm for 15 min and the resulting pellet was suspended in 25 mM Tris-HCl buffer containing 500 mM NaCl and 20 mM imidazole at pH 8.0 for lysis using high-pressure homogenizer at 4 °C. The lysate was centrifuged at 13,000 g for 30 min at 4 °C and the resulting supernatant was filtrated by 0.22 μm syringe filters for purification by Ni-NTA (Smart Lifesciences Lnc., China) and Strep-Tactin (Smart Lifesciences Lnc., China) columns. The purified protein was desalted and exchanged into 50 mM HEPES buffer containing 50 mM NaCl (pH 7.2) with an Amicon Ultra-15 centrifugal filter (10 kDa MWCO, Merk Millipore Inc.). Protein concentration was measured by Bradford assay. The obtained protein solution was added into an equal volume of glycerol for long-term storage at −80 °C.

### Blood sample measurement by NAD^+^ sensor

Phlebotomy was performed to collect venous blood samples in plastic blood collection tubes with EDTA as anticoagulant. Fingerstick sampling was performed to collect capillary blood samples using disposable fingerstick lancets (STERiLANCE Medical Inc.). The blood samples were processed by the analyzer’s automated sample handling program in which 5 μL of sample were lysed by mixing thoroughly with 4 volumes of 0.5 N perchloric acid. The lysate was then diluted by 10 folds in a buffer containing 500 mM HEPES, 500 mM NaCl at pH 7.2. 10 μL of the resulting solution was then added into 90 μL of measurement buffer containing 0.5 nM sensor, 100-fold diluted furimazine, 50 mM HEPES and 50 mM NaCl at pH 7.2, which was previously lyophilized and automatically reconstituted in the analyzer’s mixing well by the liquid handler. The sensor’s bioluminescent signal was measured using the integrated photon-detector (Hamamatsu Photonics) of the analyzer with NLuc emission measured at 460 nm (bandwidth 30 nm) and RFP emission was measured at 580 nm (bandwidth 20 nm). The sensors’ emission ratio (R) between 580 nm and 460 nm was used to calculate the NAD^+^ concentrations according to the titration curve.

To titrate the senor, standard aqueous NAD^+^ solutions with known concentrations were prepared and measured by the above-mentioned method. The sensors’ emission ratio (R) was plotted against the NAD^+^ concentrations and the resulting curve was fitted to the Hill-Langmuir equation to obtain the sensor’s maximum ratio (Rmax), minimum ratio (Rmin), c50 and Hill coefficient (h)

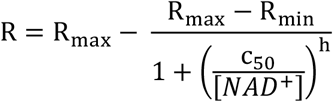

NAD^+^ concentrations in unknown samples were calculated with the rearranged the equation:

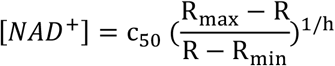

### Blood sample measurement by HPLC-MS

5 μL of whole blood was taken from blood collection tubes and added into 20 μL of 0.5 N perchloric acid with thorough mixing. Cell debris and aggregates were separated by centrifugation at 12000 g for 5 min at 4 °C. The supernatant was transferred into sample tubes and stored at −80 °C until analysis. Upon analysis, the supernatant was diluted by 10 folds in aqueous solution with 50% methanol and 50% water and then filtered with a PTFE syringe filter (0.45 μm * 13 mm) before injection into LC-MS.

Agilent Infinity Lab LC/MSD iQ G6160A was used to perform the LC-MS quantification of the blood samples. Sciex’s MultiQuant v.3.0.2 software was used for instrument control, data acquisition and data processing. 10 μL of each prepared sample were injected into LC and separated by a ZORBAX SB-Aq Column (Agilent, 4.6 × 100 mm, 3.5 μm). The autosampler temperature was set to 4 °C and the column temperature was set to 20 °C. The following elution program was used: A = acetonitrile, B = 0.1% formic acid-methyl alcohol, 0-0.3 min, 80% A; 0.3-1.5 min, 80-20% A; 1.5-2.5 min, 20% A; 2.5-2.6 min, 20-80% A; 2.6-3.0 min, 80% A, with total run time of 3.0 min and a flow rate of 1.00 mL/min. For MS quantification, MS was equipped with electrospray ionization (ESI) ion source and operates in negative ion mode. Standard solutions with known concentrations of NAD^+^ (0.05, 0.1, 0.2, 0.5, 1.0, 2.0 μg/mL) have been prepared and automatically injected with single injections for LC-MS analysis to optimize the quantification conditions. Precursor ions with the highest response were used as quantitative ion pairs and daughter ions with the higher abundance and less interference were used as qualitative pairs. The data analysis was performed with Sciex’s MultiQuant v.3.0.2 software. This quantification method showed a limit of detection (Signal to noise ratio ≥ 3) of 0.0200 μg/mL and a limit of quantification (Signal to noise ratio ≥ 10) of 0.0500 μg/mL.

| Analyte | Ion pairs (m/z) | Cataclastic voltage (V) | Dry temperature ($^{\circ}\text{C}$) | Ion source atomizing gas (psi) | Capillary voltage (V) | Retention time (min) |
| --- | --- | --- | --- | --- | --- | --- |
| $\text{NAD}^+$ | 662.0*/539.7 | 100/100 | 325 | 35 | 3500 | 1.1 |
|  | 662.0*/80.0 | 100/100 |  |  |  | 1.1 |

\*Quantitative ion pair

### Determination of recovery rate and sample stability

100 μL of solution was taken from lysed blood samples with known NAD^+^ concentrations. The samples were then added with 100, 400 and 700 μg/L standard NAD^+^. The samples were further diluted in aqueous solution with 50% methanol and 50% water for six parallel LC-MS measurements.

Blood samples lysed with 0.5 N perchloric acid were stored at room temperature and at 4 °C for 0 h, 1 h, 2 h, 4 h, 6 h, 12 h, 24 h, and 48h before LC-MS measurement using the above-mentioned method.

### Salivary sample measurement

Salivary samples were collected in 10 mL conical tubes in the morning before breakfast after at least 8 hours of fasting, which were then added with various concentrations of NAD^+^ solutions to prepare spiked salivary samples. The resulting samples were loaded into the NAD^+^ analyzer and were automatically added with 4 volumes of 0.5 N HClO_4_. The analyzer then dilutes the resulting mixture by 10 folds in a buffer containing 500 mM HEPES, 500 mM NaCl at pH 7.2, and transfers 10 μL of the resulting solution into 90 μL of measurement buffer containing 0.5 nM sensor, 100-fold diluted furimazine, 50 mM HEPES, 50 mM NaCl at pH 7.2 for bioluminescent measurement. The sensor’s bioluminescent signal was measured using the analyzer’s integrated photon-detector (Hamamatsu Photonics) with NLuc emission measured at 460 nm (bandwidth 30 nm) and RFP emission was measured at 580 nm (bandwidth 20 nm). The sensors’ emission ratio (R) between 580 nm and 460 nm was used to calculate the NAD^+^ concentrations according to the titration curve performed in parallel. HPLC-MS measurement of the supernatant was performed in parallel according to the above-mentioned method as reference.

### Statistics

All values were expressed as mean ± standard deviation. Statistics were performed using Prism 8 (Graphpad). All quantification tests were independently performed for at least 3 times.

**Correspondence and requests for materials** should be addressed to L.C., M.H. and Q.Y..

## Data Availability

All data produced in the present study are available upon reasonable request to the authors

## ACKNOWLEDGEMENT

This work is supported by Shenzhen Science and Technology Program (ZDSYS20210623091810032), National Key R&D Program of China (2020YFC2002904, 2021YFF1200302), Research Projects on Key Areas of General Colleges and Universities, Department of Education of Guangdong Province (2022ZDZX2071) Shenzhen Institute of Advanced Technology, Chinese Academy of Sciences, and Celfull Hebe Fund for the Biological Study of Aging.

## AUTHOR CONTRIBUTIONS

Q.Y., M.H., and L.C. conceived the study. L.C. and P.W. prepared the sensor. P.W., L.C., and Y.H. performed sensor qualifications. M.C. performed HPLC-MS quantifications. M.H. and J.L. provided clinical samples. R.L. contributed to the sample collection. All authors contributed to data analysis or manuscript writing.

## Reference

(1) Katsyuba, E.; Romani, M.; Hofer, D.; Auwerx, J. NAD+ Homeostasis in Health and Disease. Nat Metab 2020, 2 (1), 9–31. https://doi.org/10.1038/s42255-019-0161-5.

(2) Imai, S.; Guarente, L. NAD+ and Sirtuins in Aging and Disease. Trends in Cell Biology 2014, 24 (8), 464–471. https://doi.org/10.1016/j.tcb.2014.04.002.

(3) Chini, C. C. S.; Zeidler, J. D.; Kashyap, S.; Warner, G.; Chini, E. N. Evolving Concepts in NAD+ Metabolism. Cell Metabolism 2021, 33 (6), 1076–1087. https://doi.org/10.1016/j.cmet.2021.04.003.

(4) Covarrubias, A. J.; Perrone, R.; Grozio, A.; Verdin, E. NAD+ Metabolism and Its Roles in Cellular Processes during Ageing. Nat Rev Mol Cell Biol 2021, 22 (2), 119–141. https://doi.org/10.1038/s41580-020-00313-x.

(5) Rajman, L.; Chwalek, K.; Sinclair, D. A. Therapeutic Potential of NAD-Boosting Molecules: The In Vivo Evidence. Cell Metabolism 2018, 27 (3), 529–547. https://doi.org/10.1016/j.cmet.2018.02.011.

(6) Kumar, R.; Mohan, N.; Upadhyay, A. D.; Singh, A. P.; Sahu, V.; Dwivedi, S.; Dey, A. B.; Dey, S. Identification of Serum Sirtuins as Novel Noninvasive Protein Markers for Frailty. Aging Cell 2014, 13 (6), 975–980. https://doi.org/10.1111/acel.12260.

(7) Weiqian, C.; Caihong, Y.; Jin, L. The Role of Nicotinamide Adenine Dinucleotide in the Pathogenesis of Rheumatoid Arthritis: Potential Implications for Treatment. European Medical Journal 2018.

(8) Busso, N.; Karababa, M.; Nobile, M.; Rolaz, A.; Gool, F. V.; Galli, M.; Leo, O.; So, A.; Smedt, T. D. Pharmacological Inhibition of Nicotinamide Phosphoribosyltransferase/Visfatin Enzymatic Activity Identifies a New Inflammatory Pathway Linked to NAD. PLOS ONE 2008, 3 (5), e2267. https://doi.org/10.1371/journal.pone.0002267.

(9) Yuan, Y.; Liang, B.; Liu, X.-L.; Liu, W.-J.; Huang, B.-H.; Yang, S.-B.; Gao, Y.-Z.; Meng, J.-S.; Li, M.-J.; Ye, T.; Wang, C.-Z.; Hu, X.-K.; Xing, D.-M. Targeting NAD+: Is It a Common Strategy to Delay Heart Aging? Cell Death Discov. 2022, 8 (1), 230. https://doi.org/10.1038/s41420-022-01031-3.

(10) Carpenter, B. J.; Dierickx, P. Circadian Cardiac NAD+ Metabolism, from Transcriptional Regulation to Healthy Aging. American Journal of Physiology-Cell Physiology 2022. https://doi.org/10.1152/ajpcell.00239.2022.

(11) Imai, S.; Armstrong, C. M.; Kaeberlein, M.; Guarente, L. Transcriptional Silencing and Longevity Protein Sir2 Is an NAD-Dependent Histone Deacetylase. Nature 2000, 403 (6771), 795–800. https://doi.org/10.1038/35001622.

(12) Pirinen, E.; Auranen, M.; Khan, N. A.; Brilhante, V.; Urho, N.; Pessia, A.; Hakkarainen, A.; Kuula, J.; Heinonen, U.; Schmidt, M. S.; Haimilahti, K.; Piirilä, P.; Lundbom, N.; Taskinen, M.-R.; Brenner, C.; Velagapudi, V.; Pietiläinen, K. H.; Suomalainen, A. Niacin Cures Systemic NAD+ Deficiency and Improves Muscle Performance in Adult-Onset Mitochondrial Myopathy. Cell Metabolism 2020, 31 (6), 1078-1090.e5. https://doi.org/10.1016/j.cmet.2020.04.008.

(13) Yoshino, J.; Baur, J. A.; Imai, S. NAD+ Intermediates: The Biology and Therapeutic Potential of NMN and NR. Cell Metabolism 2018, 27 (3), 513–528. https://doi.org/10.1016/j.cmet.2017.11.002.

(14) Khan, N. A.; Auranen, M.; Paetau, I.; Pirinen, E.; Euro, L.; Forsström, S.; Pasila, L.; Velagapudi, V.; Carroll, C. J.; Auwerx, J.; Suomalainen, A. Effective Treatment of Mitochondrial Myopathy by Nicotinamide Riboside, a Vitamin B3. EMBO Molecular Medicine 2014, 6 (6), 721–731. https://doi.org/10.1002/emmm.201403943.

(15) Chini, E. N.; Chini, C. C. S.; Netto, J. M. E.; Oliveira, G. C. de; Schooten, W. van. The Pharmacology of CD38/NADase: An Emerging Target in Cancer and Diseases of Aging. Trends in Pharmacological Sciences 2018, 39 (4), 424–436. https://doi.org/10.1016/j.tips.2018.02.001.

(16) Goody, M. F.; Henry, C. A. A Need for NAD+ in Muscle Development, Homeostasis, and Aging. Skeletal Muscle 2018, 8 (1), 9. https://doi.org/10.1186/s13395-018-0154-1.

(17) Yoshino, M.; Yoshino, J.; Kayser, B. D.; Patti, G. J.; Franczyk, M. P.; Mills, K. F.; Sindelar, M.; Pietka, T.; Patterson, B. W.; Imai, S.-I.; Klein, S. Nicotinamide Mononucleotide Increases Muscle Insulin Sensitivity in Prediabetic Women. Science 2021, 372 (6547), 1224–1229. https://doi.org/10.1126/science.abe9985.

(18) Yang, F.; Deng, X.; Yu, Y.; Luo, L.; Chen, X.; Zheng, J.; Qiu, Y.; Xiao, F.; Xie, X.; Zhao, Y.; Guo, J.; Hu, F.; Zhang, X.; Ju, Z.; Zhou, Y. Association of Human Whole Blood NAD+ Contents With Aging. Frontiers in Endocrinology 2022, 13.

(19) Balashova, N. V.; Zavileyskiy, L. G.; Artiukhov, A. V.; Shaposhnikov, L. A.; Sidorova, O. P.; Tishkov, V. I.; Tramonti, A.; Pometun, A. A.; Bunik, V. I. Efficient Assay and Marker Significance of NAD+ in Human Blood. Front Med (Lausanne) 2022, 9, 886485. https://doi.org/10.3389/fmed.2022.886485.

(20) Yoshino, J.; Imai, S. Accurate Measurement of Nicotinamide Adenine Dinucleotide (NAD+) with High-Performance Liquid Chromatography. In Sirtuins: Methods and Protocols; Hirschey, M. D., Ed.; Methods in Molecular Biology; Humana Press: Totowa, NJ, 2013; pp 203–215. https://doi.org/10.1007/978-1-62703-637-5_14.

(21) Klaidman, L. K.; Leung, A. C.; Adams, J. D. High-Performance Liquid Chromatography Analysis of Oxidized and Reduced Pyridine Dinucleotides in Specific Brain Regions. Analytical Biochemistry 1995, 228 (2), 312–317. https://doi.org/10.1006/abio.1995.1356.

(22) Trammell, S. A.; Brenner, C. TARGETED, LCMS-BASED METABOLOMICS FOR QUANTITATIVE MEASUREMENT OF NAD+ METABOLITES. Computational and Structural Biotechnology Journal 2013, 4 (5), e201301012. https://doi.org/10.5936/csbj.201301012.

(23) Bustamante, S.; Jayasena, T.; Richani, D.; Gilchrist, R. B.; Wu, L. E.; Sinclair, D. A.; Sachdev, P. S.; Braidy, N. Quantifying the Cellular NAD+ Metabolome Using a Tandem Liquid Chromatography Mass Spectrometry Approach. Metabolomics 2017, 14 (1), 15. https://doi.org/10.1007/s11306-017-1310-z.

(24) Giner, M. P.; Christen, S.; Bartova, S.; Makarov, M. V.; Migaud, M. E.; Canto, C.; Moco, S. A Method to Monitor the NAD+ Metabolome—From Mechanistic to Clinical Applications. International Journal of Molecular Sciences 2021, 22 (19), 10598. https://doi.org/10.3390/ijms221910598.

(25) Graeff, R.; Lee, H. C. A Novel Cycling Assay for Cellular CADP-Ribose with Nanomolar Sensitivity. Biochemical Journal 2002, 361 (2), 379–384. https://doi.org/10.1042/bj3610379.

(26) Shabalin, K.; Nerinovski, K.; Yakimov, A.; Kulikova, V.; Svetlova, M.; Solovjeva, L.; Khodorkovskiy, M.; Gambaryan, S.; Cunningham, R.; Migaud, M. E.; Ziegler, M.; Nikiforov, A. NAD Metabolome Analysis in Human Cells Using 1H NMR Spectroscopy. International Journal of Molecular Sciences 2018, 19 (12), 3906. https://doi.org/10.3390/ijms19123906.

(27) Lu, M.; Zhu, X.-H.; Chen, W. In Vivo 31P MRS Assessment of Intracellular NAD Metabolites and NAD+/NADH Redox State in Human Brain at 4 T. NMR in Biomedicine 2016, 29 (7), 1010–1017. https://doi.org/10.1002/nbm.3559.

(28) de Graaf, R. A.; Behar, K. L. Detection of Cerebral NAD+ by in Vivo 1H NMR Spectroscopy. NMR in Biomedicine 2014, 27 (7), 802–809. https://doi.org/10.1002/nbm.3121.

(29) Yu, Q.; Pourmandi, N.; Xue, L.; Gondrand, C.; Fabritz, S.; Bardy, D.; Patiny, L.; Katsyuba, E.; Auwerx, J.; Johnsson, K. A Biosensor for Measuring NAD+ Levels at the Point of Care. Nat Metab 2019, 1 (12), 1219–1225. https://doi.org/10.1038/s42255-019-0151-7.

(30) Chen, L.; Chen, M.; Luo, M.; Li, Y.; Liao, B.; Hu, M.; Yu, Q. Ratiometric NAD+ Sensors Reveal Subcellular NAD+ Modulators. bioRxiv May 20, 2022, p 2022.05.20.491061. https://doi.org/10.1101/2022.05.20.491061.

(31) Teruya, T.; Goga, H.; Yanagida, M. Human Age-Declined Saliva Metabolic Markers Determined by LC–MS. Sci Rep 2021, 11 (1), 18135. https://doi.org/10.1038/s41598-021-97623-7.

(32) Lautrup, S.; Sinclair, D. A.; Mattson, M. P.; Fang, E. F. NAD+ in Brain Aging and Neurodegenerative Disorders. Cell Metabolism 2019, 30 (4), 630–655. https://doi.org/10.1016/j.cmet.2019.09.001.

(33) Airhart, S. E.; Shireman, L. M.; Risler, L. J.; Anderson, G. D.; Nagana Gowda, G. A.; Raftery, D.; Tian, R.; Shen, D. D.; O’Brien, K. D. An Open-Label, Non-Randomized Study of the Pharmacokinetics of the Nutritional Supplement Nicotinamide Riboside (NR) and Its Effects on Blood NAD+ Levels in Healthy Volunteers. PLoS One 2017, 12 (12), e0186459. https://doi.org/10.1371/journal.pone.0186459.

(34) Vreones, M.; Mustapic, M.; Moaddel, R.; Pucha, K. A.; Lovett, J.; Seals, D. R.; Kapogiannis, D.; Martens, C. R. Oral Nicotinamide Riboside Raises NAD+ and Lowers Biomarkers of Neurodegenerative Pathology in Plasma Extracellular Vesicles Enriched for Neuronal Origin. Aging Cell n/a (n/a), e13754. https://doi.org/10.1111/acel.13754.

(35) Conze, D.; Brenner, C.; Kruger, C. L. Safety and Metabolism of Long-Term Administration of NIAGEN (Nicotinamide Riboside Chloride) in a Randomized, Double-Blind, Placebo-Controlled Clinical Trial of Healthy Overweight Adults. Sci Rep 2019, 9 (1), 9772. https://doi.org/10.1038/s41598-019-46120-z.

(36) Okabe, K.; Yaku, K.; Uchida, Y.; Fukamizu, Y.; Sato, T.; Sakurai, T.; Tobe, K.; Nakagawa, T. Oral Administration of Nicotinamide Mononucleotide Is Safe and Efficiently Increases Blood Nicotinamide Adenine Dinucleotide Levels in Healthy Subjects. Front Nutr 2022, 9, 868640. https://doi.org/10.3389/fnut.2022.868640.

(37) McReynolds, M. R.; Chellappa, K.; Chiles, E.; Jankowski, C.; Shen, Y.; Chen, L.; Descamps, H. C.; Mukherjee, S.; Bhat, Y. R.; Lingala, S. R.; Chu, Q.; Botolin, P.; Hayat, F.; Doke, T.; Susztak, K.; Thaiss, C. A.; Lu, W.; Migaud, M. E.; Su, X.; Rabinowitz, J. D.; Baur, J. A. NAD+ Flux Is Maintained in Aged Mice despite Lower Tissue Concentrations. Cell Systems 2021, 12 (12), 1160-1172.e4. https://doi.org/10.1016/j.cels.2021.09.001.

(38) McReynolds, M. R. NAD+ Flux and Resiliency in Aged Mice. The FASEB Journal 2022, 36 (S1). https://doi.org/10.1096/fasebj.2022.36.S1.R6281.

