## supplemental material for "Fingerstick blood assay maps real-world NAD+ disparity across gender and age"

**Table S1. Age and gender information of subjects for evaluating NMN supplementation without sport.**

|  | Placebo | 500 mg NMN | 1000 mg NMN |
| --- | --- | --- | --- |
| Sample size | 25 | 19 | 21 |
| Gender (M/F) | 10/15 | 9/10 | 10/11 |
| Average age | 57.00 ± 6.86 | 57.95 ± 5.23 | 58.10 ± 4.22 |
| Sport (Yes/No) | No | No | No |
| Average NAD^+^ | 23.8 ± 5.5 | 41.7 ± 13.0 | 58.8 ± 21.1 |

**Table S2. Age and gender information of subjects for evaluating NMN supplementation in combination with moderate level of sport.**

|  | Placebo | 500 mg NMN |
| --- | --- | --- |
| Sample size | 21 | 20 |
| Gender (M/F) | 10/11 | 10/10 |
| Average age | 58.21 ± 5.39 | 58.59 ± 4.75 |
| Sport (Yes/No) | Yes | Yes |
| Average NAD^+^ | 33.18 ± 7.18 | 55.48 ± 21.37 |

**Table S3. Age and gender information of subjects for surveying NAD^+^ level via fingerstick sample**

| Age range | 18-50 | | 50-85 | |
| --- | --- | --- | --- | --- |
| Gender | M | F | M | F |
| Sample size | 24 | 23 | 46 | 56 |
| Average age | 33.42 ± 7.02 | 33.35 ± 8.56 | 67.88 ± 9.33 | 67.23 ± 8.55 |
| Average NAD^+^ | 44.2 ± 18.9 | 32.7 ± 9.6 | 25.9 ± 9.8 | 24.8 ± 9.6 |

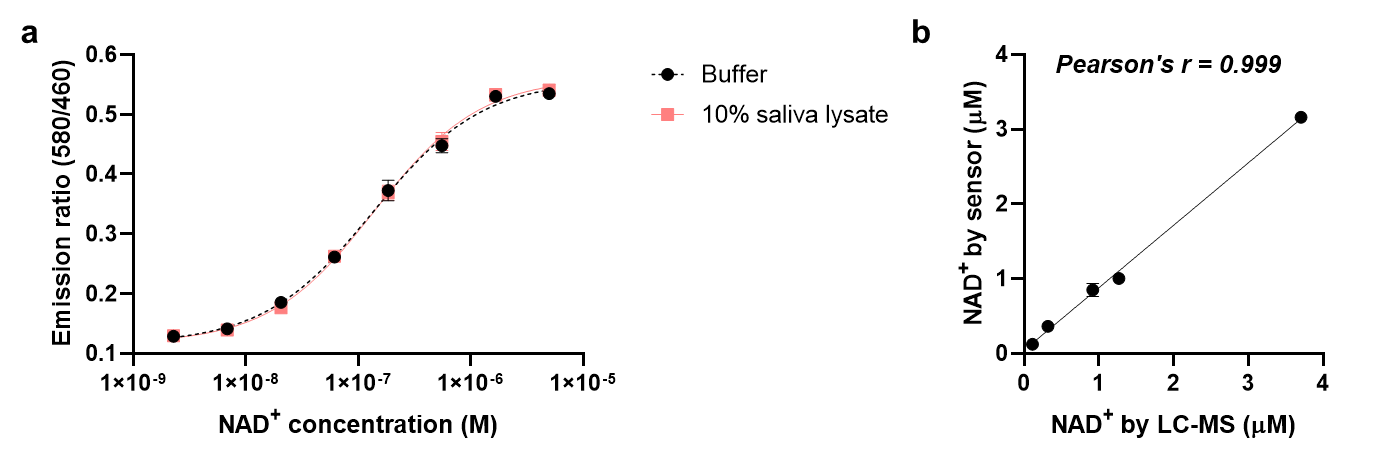

**Figure S1. Measurement of NAD^+^ in spiked saliva samples. (a)** Sensor titration curve in standard buffer and buffer spiked with 10% saliva lysate. The presence of saliva lysate did not show interference with the sensor performance. **(b)** Comparison between sensor and LC-MS measurement of NAD^+^ in saliva. The two methods showed a high level of agreement with Pearson’s r = 0.999. In (a) and (b), values are given as mean ± SD of three independent measurements.

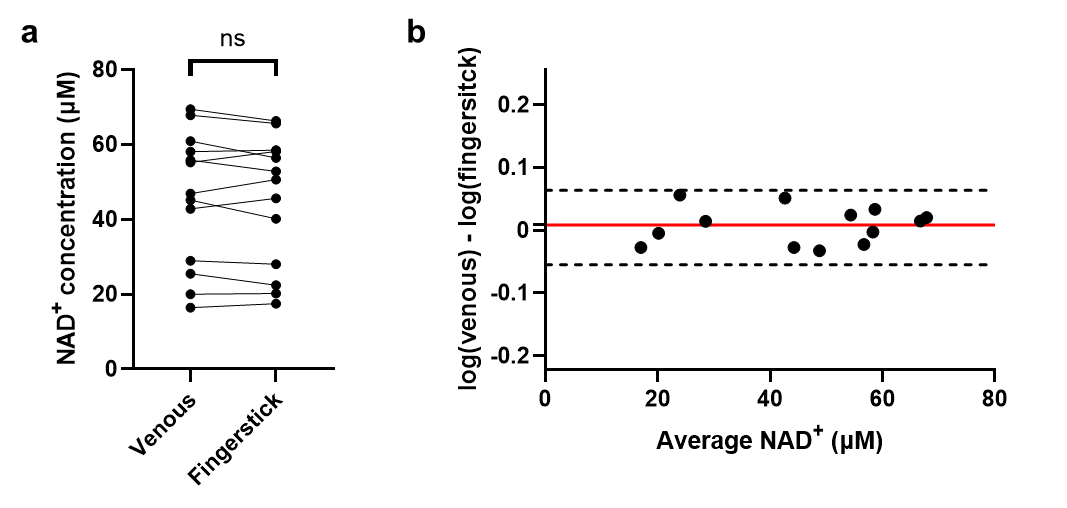
**Figure S2. Correlation between fingerstick and venous NAD^+^. (a)** Comparison of NAD^+^ levels measured from venous and fingerstick samples. No significant difference was found between the two sample types. Significance was determined using paired t-test. **(b)** Bland-Altman analysis for venous NAD^+^ measured from venous and fingerstick blood samples.

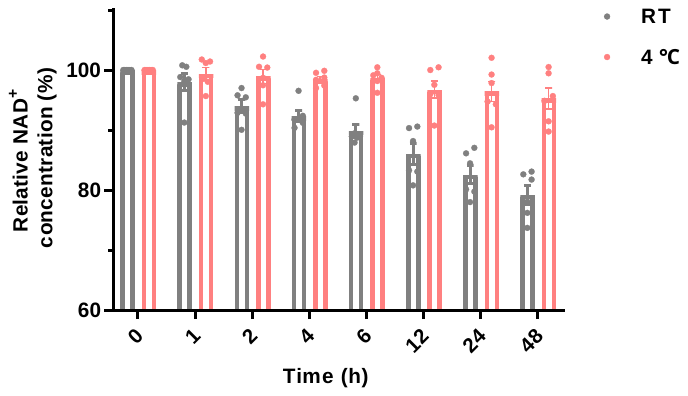
**Figure S3. Degradation of NAD^+^ in biological samples at room temperature and 4 ^◦^C measured by LC-MS.** Storage at room temperature induced considerable degradation of NAD^+^ in blood samples compared to the storage at 4 ^◦^C. Error bars represent SD of n = 6 independent biological repeats.
